# Antibodies to SARS/CoV-2 in arbitrarily-selected Atlanta residents

**DOI:** 10.1101/2020.05.01.20087478

**Authors:** Jun Zou, Alexis Bretin, Andrew T. Gewirtz

## Abstract

We quantitated anti-SARS/CoV-2 IgG and IgM by ELISA in self-collected blood samples (n=142) in arbitrarily-selected metro Atlanta residents, primarily acquaintances of the authors’ lab members from 4/17-4/27, 2020. Archived serum (n=34), serum from nucleic acid test (NAT)-positive subjects (n=4), and samples collected from NAT-positive community members (n=4) served to validate the assay. The range of anti-SARS/CoV-2 antibodies in archived and NAT-positive sera indicated need to compromise sensitivity or specificity. Accordingly, we set a cutoff of 4 SD above the mean for IgG and 3 SD above the mean for IgM to indicate that an individual had been exposed, and developed some degree of immunity, to SARS/CoV-2. The IgG cutoff clearly compromised sensitivity but offered high specificity, both of which were harder to gauge for IgM. Based on these cutoffs, excluding subjects whose participation resulted from self-suspected SARS/CoV-2 infection, we found 7.1% positivity for anti-SARS/CoV-2 IgG (3 of 127 subjects) or IgM (6 of 127). While we do not claim this small immune survey is broadly representative of metro Atlanta, and we have greater confidence in the IgG results, which had only 2.4% positivity, it nonetheless demonstrates that persons with antibodies to SARS/CoV-2, who’ve not suspected they’d been exposed to this virus, can readily be found in various Atlanta area neighborhoods (9 positives were in 8 zip codes). Accordingly, these results support the notion that dissemination of the virus is more widespread than testing would indicate but also suggests that most persons in metro Atlanta remain vulnerable to this virus. More generally, these results support the general utility of sero-surveillance to guide public policy but also highlight the difficulty of discerning if individuals have immunity to SARS/CoV-2.

## Introduction

There is now a wide interest in measuring SARS-CoV-2 antibodies both as a means of gauging the true extent to which persons within a particular geographic region have been infected by this virus and perhaps as a means of discerning whether individuals might now be immune from SARS/CoV-2-induced disease. To address the former, some have begun conducting “sero-surveys”. One recently publicly-posted study described use of a lateral flow test, which purports to give a qualitative yes or no for presence of absence of such antibodies and observed that 4.3 *%* of tested resident of Santa Clara CA had been exposed to the virus [1]. Press releases of other studies that did not give details of their methodology observed presence of SARS-CoV-2 antibodies in 14.3% of resident in Heinsberg, Germany and 18% of residents in New York State. Herein, we report results of our effort to measure SARS/CoV-2 antibodies in resident of metropolitan Atlanta, GA. We analyzed self-collected blood samples from acquaintances of the authors’ lab members by a quantitative lab-based ELISA method. We found that antibodies to SARS/CoV-2 are not merely a yes or no but have a broad quantitative range, with some overlap when comparing samples from confirmed infected subjects versus samples collected before the advent of SARS-CoV-2. Nonetheless, our quantitative approaches enabled seemingly readily and reliable discernment that about 3 and 6 of 127 arbitrarily subjected Atlanta area residents displayed SARs/CoV-2-specific IgG and IgM, respectively, which likely reflects that they have been exposed to this virus.

## Methods

All procedures received required approvals by Georgia State University’s Institutional Review Board and/or Biosafety committee.

### Subject selection

An advertisement was prepared that explained that the goal of the study was to perform community by sero-surveillance, and specified that no results would be provided to individual participants. The advertisement was circulated by the authors and their lab members in their neighborhoods (see table 1 for subject zip codes), which resulted in enrollment of 142 individuals, 135 of whom had not been tested for SARS/CoV-2 and 2 individuals whom had suspected they’d been infected but tested negative at an area hospital. While these subjects were not asked any health-related questions, some volunteered that they suspected they might have been previously infected, which we noted. Additionally, we enrolled 4 community members whom had been referred to the study by word of mouth primarily because they had been deemed positive for SARS/CoV-2 by a PCR-based test. One of these subjects recruited 5 household members, one of whom exhibited symptoms. We also enrolled 4 subjects who had not been tested for SARS/CoV-2 but yet were referred to the study because they had told associates of the study team that they believed they had been infected by SARS/CoV-2. Additionally, 3 serum samples from NAT-positive subjects were provided by Sean Stowell (Emory University) and one was provided by Mount Sinai School of Medicine Department of Pathology.

### Blood collection and serum isolation

Subjects were provided an autoclaved 1.5 ml Eppendorf tube, 2 26-gauge safety lancets, and asked to watch a video demonstrating collection of 1-2 blood droplets https://www.youtube.com/watch?v=H9e-9xvhrms. Subjects were advised to then collect their blood and contact study team. This procedure generally resulted in blood being centrifuged (5000 rpm, 10 minutes) and serum isolated in a biosafety cabinet at GSU within 24 hours of blood collection, although test comparisons of allowing storing blood for up to 3 days did not seem to impact results. This procedure yielded over 5 and 10 microliters of serum, respectively in over 97% and 93% of subjects respectively. Serum was then heated, in sealed Eppendorf tubes, to 55°C for 15 minutes to reduces it infective potential. Archived control sera consisted of samples collected from healthy control subjects in the Atlanta area from 2015-2019 that had been stored at −80°C. Such sera was heat-inactivated as above prior to use in this study.

### ELISA

(Based on protocols from Krammer and colleagues [2])

Recombinant his-tagged S1 SARS/CoV-2 S1 Spike protein, purchased from Sino Biologicals (Wayne PA), was diluted to a concentration of 1 μg/ml in PBS and applied overnight, 100 μl per well, to Ni-coated 96-well plates (Fisher/Thermo) at 4 °C. Plates were then blocked with PBS/ 5% skim milk for 1h at room temperature, washed 3X with PBST (PBS 0.1% tween 20). Serum, sequentially diluted 1:100 in PBS/1% BSA and 1: 10 in 3% skim milk respectively, was then applied to wells for 1h at 37 °C. Plates were then washed 3X with PBST and then incubated with antihuman IgG or IgM (KPL), which was diluted 1:5000 and 1:50,000 respectively. 45 min later, plates were washed 3X with PBST and developed with TMB for 10 minutes at which time reaction was stopped with stop reagent (KPL, TMB Stop solution) and OD450 nm measured by a 96-well plate reader (subtracting readings at 540 nm). A positive control for the assay was used to assess and correct for plate to plate variance of the assay. In the course, of setting and validating this assay, many samples were assayed multiple times and gave similar relative patterns of results although absolute OD varied between assays run on different days. Hence, all data reported herein results from all samples being assayed together.

## Results

Some of the archived serum samples, which were collected well before SARS/CoV-2 is thought to have existed, displayed modest but measurable immune reactivity to recombinant SARS/CoV-2 S1 spike protein (Figure 1), likely reflecting non-specific binding or cross-reactivity to other Coronavirus spike proteins. In any case, we presumed that any newly collected serum samples displaying immune reactivity to SARS/CoV-2 spike protein above the range of the archived samples likely reflects that an individual has been exposed to SARS/CoV-2. Conversely, test samples having values within the range of the archived samples reflects an individual’s lack of exposure to SARS/CoV-2 or that the antibody response an individual generated at the time of blood collection was not sufficient to rise above the range of the archived samples. Indeed, one study reported that 30% of hospitalized subjects don’t make antibodies to the virus [3]. For example, one of the 4 hospital-provided NAT+ serum samples yielded an OD within the range of the control samples. Hence, we concluded that setting a specific cut-off to declare samples SARS/CoV-2 positive or negative will require compromising sensitivity or specificity. Accordingly, as all archived sample fell within 2.6 SD of the group mean, we set a cut-off of 4 SD above that mean as a threshold to indicate that an individual had likely been exposed to SARS/CoV-2. If values had a normal distribution, such a cut-off would provide over 99% specificity. This cut-off result was met by 3 of 4 community-collected NAT+ samples. The IgM assay was harder to validate in that, as one might expect, the 4 hospital-provided convalescent serum samples were all within the range of the archived samples. Hence, based on overall analysis of our data and other studies, we set a cutoff of 3 SD to indicate likely recent exposure to SARS/CoV-2. Based on this cutoff, 2 of our 4 community-acquired NAT+ samples, including the one that was negative for IgG, were positive for SARS/CoV-2 anti-IgM. Thus, all 4 community-collected subjects whom reported NAT+ to us exhibited anti-SARS/CoV-2 IgG or IgM.

**Figure 1.**
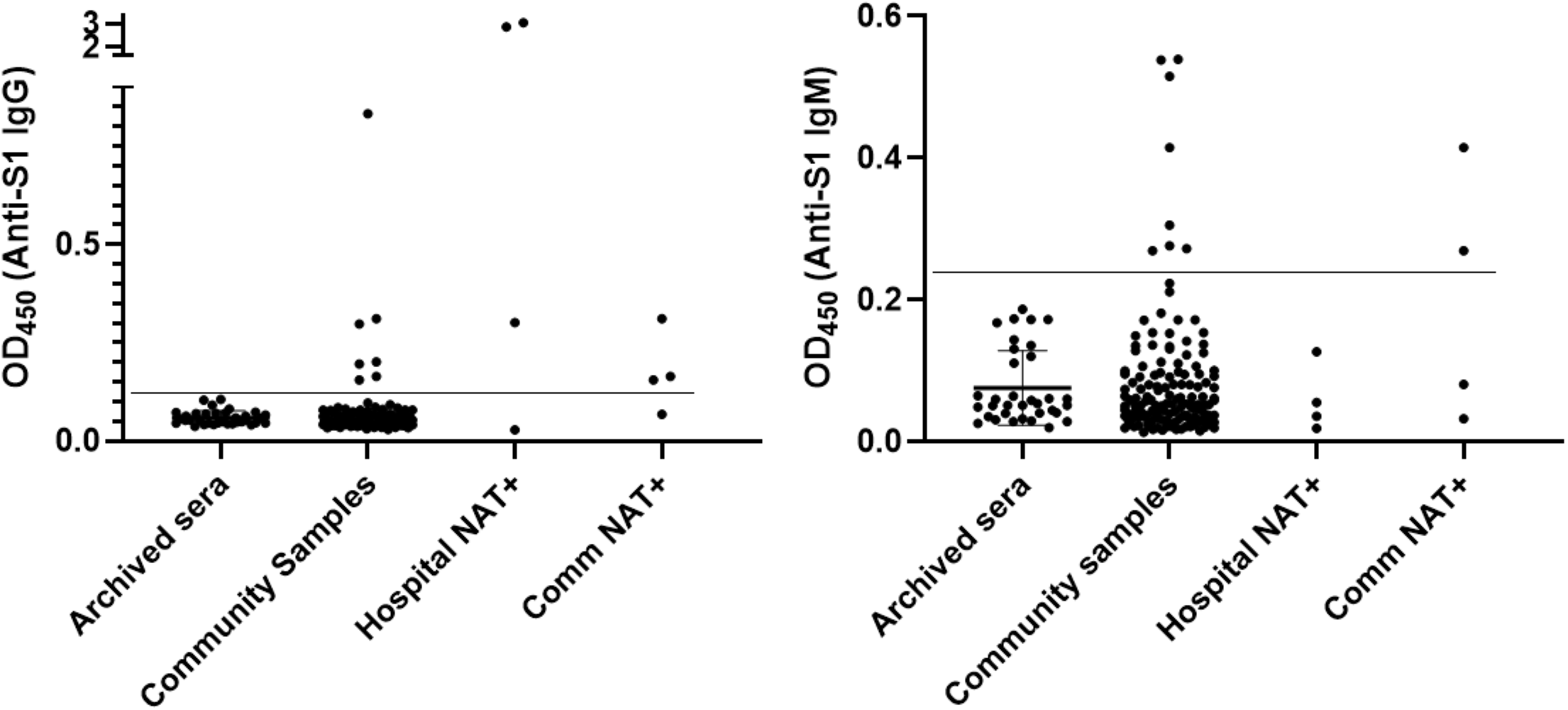
SARS/Cov-2 antibody levels by ELISA. Data generated as described in text. Horizontal line indicates the 4 and 3 SD above the mean cutoff used for IgG and IgM.

Based on the above-defined cutoffs, 7 of all 142 community-collected samples we collected in this study were positive for SARS/CoV-2 IgG. Yet, the level of SARS/CoV-2 IgG in all of these samples was dramatically lower than the 2 high-titer hospital-provided positive control samples. We speculate that severe SARS/CoV-2 infections may result in viremia that drives very high antibody responses whereas cases not requiring hospitalization result in more subtle and/or delayed antibody responses. Accordingly, none of the blood samples we tested (n=20), including 2 IgG+ and 2 IgM+ subjects, showed detectable virus in blood by PCR. Of the 7 IgG+ samples we collected from the community, 3 were from NAT+ subjects and one was a household member of one of those subject. This latter sample yielded more than double the OD of the sample provided by the overtly ill NAT+ subject despite the household member lacking severe symptoms. Several other subjects were included in the study purely due to having contacted the study team after having heard of the study second hand. Excluding both groups, results in 3 of 127 subjects that were selected based only on acquaintance to the study team as being IgG+. These positives were from 3 different zip codes (Table 1) and had no apparent connection to each other and never displayed symptoms, and hence can be viewed as unbiasedly selected to have participated in the study. Analogously, for IgM, 8 of 142 were positive of which 2 were previously found to be NAT+. Of the 127 unbiasedly-selected participants, 6 were positive, all from different zip codes. These studies support the notion that dissemination of SARS/CoV-2 is greater than confirmed testing would indicate and highlight importance of broad randomized testing for past and present indicators of the virus.

**Table 1.** Zip codes of SARS/CoV-2 antibody testing. Zip code, number of subjects tested, IgG+, and IgM+. Highlighted values refer to the positive tests from persons whose selection was not based on suspected positivity.

| zip | tested | IgG | IgM |
| --- | --- | --- | --- |
| 30030 | 10 | 0 | 0 |
| 30033 | 32 | 1 | 1 |
| 30038 | 8 | 2 | 0 |
| 30041 | 1 | 0 | 0 |
| 30075 | 13 | 0 | 1 |
| 30094 | 2 | 0 | 1 |
| 30096 | 1 | 0 | 0 |
| 30097 | 1 | 0 | 0 |
| 30135 | 4 | 1 | 0 |
| 30188 | 5 | 0 | 1 |
| 30189 | 3 | 0 | 0 |
| 30303 | 1 | 0 | 0 |
| 30305 | 3 | 1 | 1 |
| 30307 | 15 | 0 | 1 |
| 30308 | 6 | 1 | 0 |
| 30309 | 1 | 0 | 1 |
| 30312 | 3 | 0 | 0 |
| 30316 | 1 | 0 | 0 |
| 30317 | 6 | 1 | 1 |
| 30318 | 1 | 0 | 0 |
| 30319 | 3 | 0 | 0 |
| 30324 | 3 | 0 | 0 |
| 30327 | 4 | 0 | 0 |
| 30328 | 2 | 0 | 0 |
| 30329 | 4 | 0 | 0 |
| 30339 | 1 | 0 | 0 |
| 30340 | 1 | 0 | 0 |
| 30342 | 2 | 0 | 0 |
| 30344 | 1 | 0 | 0 |
| 30345 | 1 | 0 | 0 |
| 30606 | 2 | 0 | 0 |
| 31302 | 1 | 0 | 0 |

## Discussion

Determining the extent to which SARS/CoV-2 has disseminated within a particular geographic region is critical to understanding its virulence and the degree to which the community has acquired some degree of immunity to it. While testing for SARS/CoV-2, which has largely focused on those with symptoms and, as of April 27, 2020, confirmed the presence of the virus in about 0.23% of the population, true incidence of exposure is presumed to be much higher. In accord with this notion, we found that 7.1% of those we arbitrarily-selected, we submit unbiasedly, displayed levels of anti-SARS/CoV-2 antibodies that indicated likely exposure to the virus. It is difficult to determine the extent to which our survey is representative of metro Atlanta in general or even those areas that are most represented. We certainly would not claim that our selection of participants was random but yet we submit our arbitrary selection process had minimal bias in terms of whether persons suspected they’d been infected and largely resulted in inclusion of households that would not have likely had close contact in 2020. But on the other hand, our survey participants were heavily weighted toward northeast Atlanta neighborhoods and surely under-represents African Americans and Latinos, whom are thought to be most likely to be exposed to the virus. Nonetheless, that our small survey identified SARS/CoV-2 antibody-positive in subjects whom were Caucasian, African Americans, Latinos, and Asian Americans suggesting the virus has broadly disseminated in the region. The modest numbers of African Americans and Latinos included make it difficult to weight or extrapolate our results but yet attempting to so would lead us to guess that applying our methodology broadly and randomly across the region would yield higher rates of SARS/CoV-2 antibody positivity than observed in our study.

The rate of SARS/CoV-2 antibody positivity we observed is greater than the 2.4% observed in Santa Clara SC in early April and is considerably lower than that recently measured in NY. Yet, aside from our study being smaller and not having attempted random sampling, our quantitative methodology may have allowed us to detect weak but nonetheless real positives that would seem to be extremely difficult to detect by lateral flow without also picking up many false positives. Considering this presumed methodological differenced (to our knowledge methods used by NY State have not been reported in detail) and that the per capita death rate in New York State is over 10 times that of Georgia, we speculate that applying our methodology to NY would reveal higher rates of exposure than they observed by their methodology. Yet, one can readily imagine alternative explanations. For example, perhaps sequence variants of SARS/CoV-2 predominating Atlanta are less virulent than that those in NY. Perhaps the lower population density and relatively low use of public transportation in Atlanta resulted in exposures to lower doses of the virus.

During the course of our study, about 15 participants expressed the view that they strongly suspected they had been exposed to SARS/CoV-2 based on symptom; about 10 of our unbiasedly selected subjects and 5 others who contacted us based on their symptoms. Only one of these subjects displayed SARS/CoV-2 antibodies (IgM) thus indicating the difficulty of diagnosing by symptoms. Conversely, none of the 3 IgG+ positive subjects we unbiasedly identified displayed any symptoms, cautioning against presuming healthy people are not infected. Yet, that all 3 had household members lack SARS/CoV-2 antibodies fits with the notion that such individual are not “super spreaders” of the virus. In conclusion, we readily acknowledge our study faced a variety of limitations, as one might expect would be the case when a small team of mouse-based researchers seek to originate a clinical study in the course of a pandemic. Nonetheless, we believe our results can inform approaches to sero-surveil communities to better manage the pandemic.

## Data Availability

Raw data available upon request.

